# A rapid review on the utility of physical activity pre-participation questionnaires

**DOI:** 10.1101/2025.09.19.25336065

**Authors:** Elliott C.R. Hall, Ashley Ridout, Hamish Reid

## Abstract

**Background:** Exercise brings multiple health benefits and removing barriers to participation is important. Fears around adverse events during exercise, particularly those of cardiovascular nature, have traditionally underpinned conservative approaches to pre-participation clearance. Concerns exist that existing pre-participation screening questionnaires create false positives and unnecessarily inhibit people from exercising. The aims of this rapid review were (i) to describe the utility, and (ii) to assess the impact, of pre-participation screening questionnaires.

**Methods:** A literature search of the PubMed, Web of Science, Scopus, and Google Scholar databases was conducted from inception to February 2024. Included articles reported (a) background, justification, and/or commentary on development/revision/implementation of pre-participation questionnaires, (b) sensitivity, specificity, or other metrics of pre-participation questionnaires, and/or (c) stakeholder opinions regarding pre-participation questionnaire utility.

**Results:** Sixty-three reports were included, comprising original research, pre-prints, meeting abstracts, conference proceedings, literature reviews, and editorial materials. Thirteen different questionnaires were identified, and versions of the Physical Activity Readiness Questionnaire (PAR-Q, revised PAR-Q, PAR-Q+) and related follow-up tools (PARmed-X, ePARmed-X) were investigated most (61.4%). Original estimates suggested 1 in 5 respondents would be referred for medical clearance before increasing their exercise. Subsequent studies suggest this is an underestimation, especially for older individuals, and there is limited evidence supporting pre-participation questionnaires in reducing mortality and adverse events.

**Conclusions:** Pre-participation questionnaires have several limitations. False positives create unnecessary barriers to exercise, particularly in older adults, and sub-optimal test sensitivity increases the risk of “at-risk” individuals being placed at undue risk. Further work is needed to improve this process.

## 1. Introduction

### 1.1 - Background

The health benefits of regular physical activity are considerable, with the largest relative gains observed when the least fit increase their activity [1, 2]. As inactivity prevalence increases, removing barriers to exercise (a structured form of physical activity performed with the intention of maintaining or improving health) is a key objective across the globe. Fears around precipitating adverse events during exercise, particularly those of cardiovascular nature, contribute to conservative approaches to pre-participation clearance [2]. For example, in the 1970s men aged >35 in the U.S.A were recommended an exercise electrocardiogram (ECG) before becoming more active, due to the belief that medically supervised screening could predict and prevent adverse events, despite a lack of evidence indicating that this would reduce adverse events [3]. Logistics and cost rendered this approach prohibitive, prompting the development of self-administered screening protocols. Various questionnaires now signpost “at-risk” individuals to seek further clearance, either from a physician or qualified exercise professional.

The diagnostic accuracy of a screening tool lies in the ability to correctly distinguish “at-risk” individuals from those at low risk [4]. This can be determined by sensitivity and specificity, both assessed by comparing the results of a screening tool against an accepted gold-standard [5]. In the case of pre-participation questionnaires, that gold-standard is often clinical examination involving a resting and/or exercise ECG. In this context, sensitivity is the proportion of at-risk individuals identified by clinical examination who are detected by the questionnaire. Specificity is the proportion of low-risk individuals the questionnaire correctly clears to commence physical activity without requiring further investigation. A tool with high sensitivity performs well at identifying those who are contraindicated, whereas a tool with high specificity performs well at identifying those who are not [5]. There is often a trade-off between sensitivity and specificity that can lead to false positives (people being barred from physical activity who don’t need to be) or false negatives (people being referred for further medical investigation who don’t need it).

Establishing criterion validity of pre-participation questionnaires against clinical examination in sufficiently powered samples is challenging due to time and cost [3]. Consequently, the proportion of individuals responding positively to a pre-participation questionnaire can also infer whether the screening tool might generate a high number of false positives or negatives.

### 1.2 – Reasons for this review

Concerns exist regarding unnecessary barriers to exercise created by pre-participation questionnaires. This problem can lead to healthy individuals and/or those with medical conditions being unnecessarily delayed or inadvertently discouraged from exercise due to false positive screening results [6]. This can have a profound effect on confidence and motivation toward physical activity. However, almost universally, the risk of exercise-related adverse events is markedly lower than the health risk of remaining inactive [6, 7]. Nonetheless, because a legitimate risk of adverse events does exist when high risk individuals start physical activity, pre-participation screening is an important and recommended medicolegal risk management strategy for health and fitness facilities [8]. The risk of false negatives is therefore also of interest [9]. There is a need to understand the origins and validity of pre-participation questionnaires, the degree to which these tools present a barrier to exercise, and whether an exploration of alternative approaches is warranted.

### 1.3 - Aims of this review

1. Describe the utility of pre-participation questionnaires by considering:

a. Which pre-participation questionnaires are most frequently reported in published literature?
b. How were these questionnaires developed and implemented?
c. What is the diagnostic accuracy of pre-participation questionnaires?
d. What are the limitations of pre-participation questionnaires?
2. Assess the impact of pre-participation questionnaires by describing:

a. The proportion of individuals referred/advised to seek further medical clearance.
b. Any associated impact on mortality and/or adverse events

## 2. Methods

A Rapid Evidence Review was selected based on the aims of the review and due to the time and resources available. The *a priori* decision to undertake a rapid review acknowledged the possibility of incomplete search coverage and limited capacity for quality assessment, and that synthesis and analysis of evidence would principally be tabular and narrative [10]. We consulted the *“Updated recommendations for the Cochrane rapid review methods guidance for rapid reviews of effectiveness”* [11] in the design and reporting of this review. Potential limitations arising from the use of restricted methods are discussed in section 3.8

### 2.1 - Search Terms

A search for evidence was performed using four major online literature databases from inception to 28^th^ February 2024. The databases were PubMed, Web of Science, Scopus, and Google Scholar. Original search terms included: ‘pre-exercise screening questionnaire’, ‘pre-exercise health questionnaire’, ‘exercise screening questionnaire’, ‘pre-participation screening’, ‘pre-participation questionnaire’, ‘exercise health questionnaire’ and ‘exercise medical screening questionnaire’. Identification of pre-screening tools from initial search results informed additional query terms including (but not limited to): ‘physical activity readiness questionnaire’, ‘PAR-Q’, PARQ’, ‘PAR-Q+’, ‘PARmed-X’, ‘ePARmed-X’, ‘ACSM questionnaire’, ‘ACSM screening’, ‘ACSM algorithm’, ‘AHA questionnaire’, ‘SMA-AAESS’, ‘GETP9’, ‘AAPQ questionnaire’, ‘adult pre-exercise screening system’, ‘APSS’, “PESS”, ‘Get Active Questionnaire’, ‘GAQ questionnaire’, ‘ECR questionnaire’ and ‘EACPR questionnaire’.

### 2.2 - Inclusion Criteria

All document types available from the databases were considered. Titles and abstracts were screened to determine suitability against the aims of the review with full-text versions screened where available. One author (EH) performed initial screening and queries around inclusion were resolved by discussion with a second (HR). Articles not written in English were excluded, except where an English abstract was provided. Results with no abstract and no full text available were excluded. Articles where screening was not the aim of the study were included if information was available on the proportion of positive responses to pre-participation questionnaire(s). For this review, ‘positive responses’ refers to persons meeting the criteria to seek further medical clearance following completion of a pre-participation questionnaire.

Inclusion criteria to address Aim 1 “*Describe the utility of pre-participation questionnaires*”

1. Provides background, justification, and/or critical commentary on the development, implementation, and/or revision of one or more pre-participation screening questionnaires.
2. Quantifies sensitivity, specificity, or other validation metrics of pre-participation screening questionnaires.
3. Quantifies opinions of relevant stakeholders on the utility of pre-screening questionnaires.

Inclusion criteria to address Aim 2 “*Assess the impact of pre-participation questionnaires*”

4. Quantifies positive response rates and referral/advice outcomes from pre-participation screening questionnaires.
5. Quantifies risk associated with positive responses to one or more pre-screening questionnaires.
6. Evaluates the relationship between questionnaire-based screening and mortality or adverse events.

## 3. Results

### 3.1 - Summary of search results

Sixty-three reports (original research, pre-prints, meeting abstracts, conference proceedings, literature reviews, editorial materials) were identified as relevant to the aims of the review. A total of 48 reports contained quantitative (97.9%) or qualitative (2.1%) outcome measures [4, 9, 12–57]. Nine of these reports compared two or more pre-participation questionnaires in the same sample. Fifteen reports were literature reviews, consensus documents or editorial materials related to pre-participation screening [2, 3, 6, 58–69].

### 3.2 - Which pre-participation questionnaires are most frequently reported in published literature? (Aim 1a)

**Several pre-participation questionnaires are available, with variations of the Physical Activity Readiness Questionnaire (PAR-Q) reported most in published literature (Fig. 1)**

In published reports with outcome measures, versions of the Physical Activity Readiness Questionnaire (PAR-Q, revised PAR-Q, PAR-Q+) and related follow-up tools (PARmed-X, ePARmed-X) were most commonly investigated questionnaires (61.4%). Other screening tools included versions of the American College of Sports Medicine (ACSM) screening guidelines (8.8%), the American Heart Association/American College of Sports Medicine Preparticipation Questionnaire (AAPQ, 5.3%), European Association of Cardiovascular Prevention and Rehabilitation (EACPR) guidelines (5.3%), the American Heart Association questionnaire (AHA, 5.3%), European Society of Cardiology (ECR) guidelines (3.5%), the Canadian Society of Exercise Physiology (CSEP) Get Active Questionnaire (GAQ, 3.5%), the Sports Medicine Australia-Australian Association for Exercise and Sport Science screening system (SMA-AAESS, 1.8%), the Sports Medicine Australia Adult Pre-Exercise Screening System (PESS, 1.8%), the Cornell Medical Index (1.8%), and the Framingham Risk Score (1.8%).

**Fig. 1.**
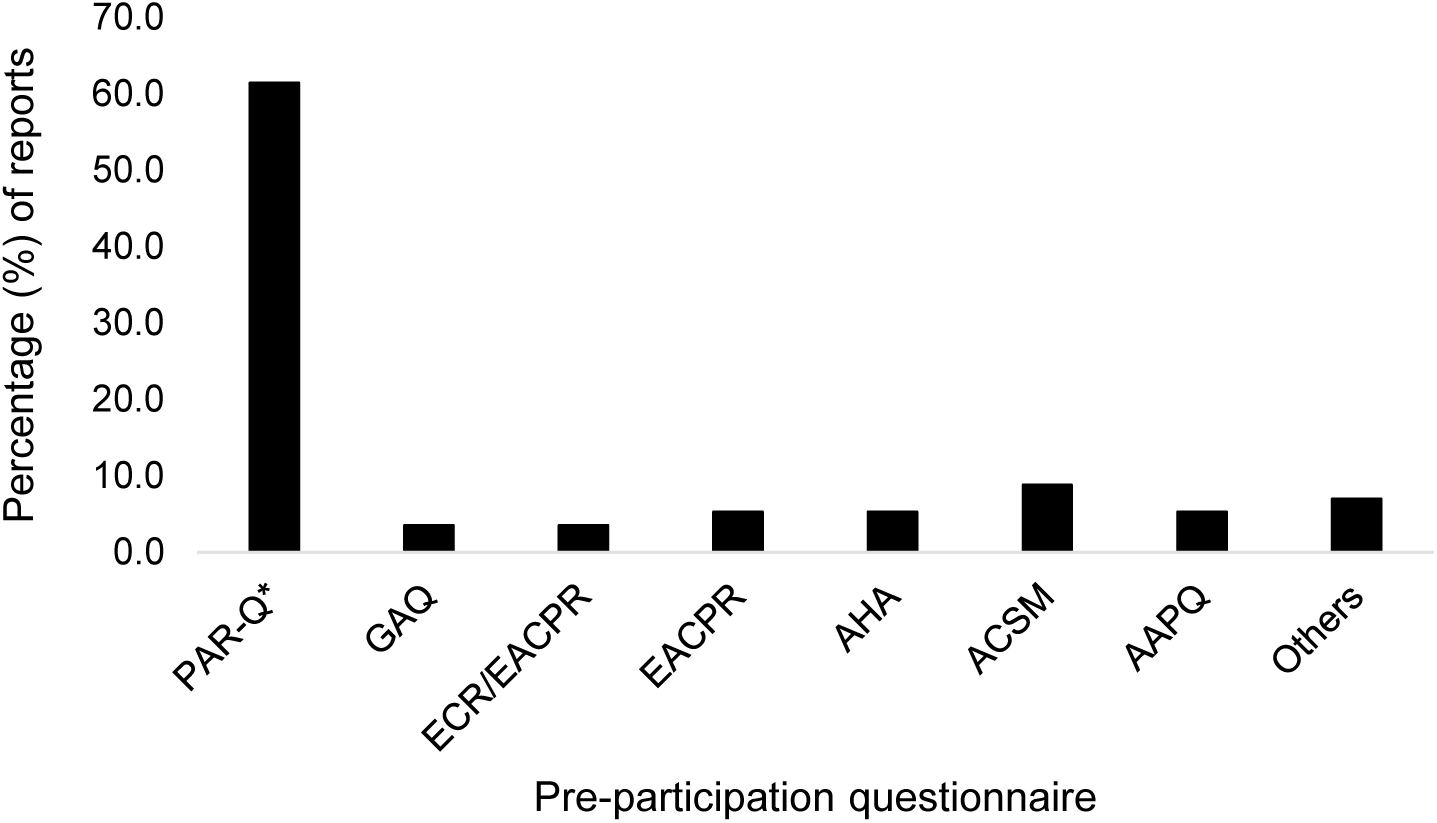
– Pre-participation questionnaires identified by rapid review process. *PAR-Q includes revised versions and PAR-Q+/ePARmed-X. **Others are SMA-AAESS, PESS, Cornell Medical Index, and Framingham Risk Score.

### 3.3 - How was the PAR-Q developed and introduced? (Aim 1b)

**The PAR-Q was developed in Canada in the 1980s and became the international standard pre-participation screening tool in the 1990s and 2000s. Revisions and updates have followed**

Reports on PAR-Q variations account for nearly two thirds of research articles identified. The PAR-Q was developed in the 1970s by the Ministry of Health in British Columbia, Canada. An eight-item self-report questionnaire was tested on 3,000 adults when validating the Canadian Home Fitness Test [14]. Originally, the questionnaire was designed to establish safety to perform the test unsupervised. An expanded nineteen-item version was validated against clinical examination including blood pressure, resting ECG and exercise ECG in 1,253 conference attendees, before expert consensus selected the 7 most relevant “Yes/No” items [70]. This 7-item tool became the PAR-Q and was accompanied by the 4-page ‘PARmed-X’, an “aide memoire” to help physicians determine whether to provide exercise clearance to persons responding “Yes” to one or more PAR-Q items [22]. Further work in the 1980s reported PAR-Q sensitivity and specificity against clinical examination (section 3.3, Table 1)[49, 50]. Initial reports suggested that the 7-item PAR-Q achieved its original aim, reducing the number of required physician clearances by as much as 90% [6] and was subsequently endorsed by governing bodies in the U.K, U.S.A and Canada in the 1990s and 2000s. The PAR-Q became the international standard pre-participation screening tool [6].

**Table 1.** – Sensitivity and specificity values from studies investigating the accuracy of pre-participation questionnaires.

| Screening Tool | Country (Setting) | Sample | Age | Comparator | Sensitivity | Specificity | Reference |
| --- | --- | --- | --- | --- | --- | --- | --- |
| PAR-Q<br>(19-item version) | Canada<br>(Agricultural exhibition) | 1,253 | Adults<br>(unspecified) | Clinical exam, BP | 92.5 | 78.5 | [22] |
|  |  |  |  | Clinical exam, BP, resting ECG | 91.6 | 72.9 |  |
|  |  |  |  | Clinical exam, BP, resting ECG, stress ECG | 41.8 | 41.8 |  |
| PAR-Q | Canada<br>(Insurance employees) | 1,130 | 18 - 70 | Clinical examination | 100.0 | 81.5 | [49] |
|  |  |  |  | Stress ECG | 33.3 | 82.1 |  |
| PAR-Q | Canada<br>(University staff) | 74 | 45-75 | Clinical exam | 55.0 | 83.3 | [50] |
| PAR-Q (revised) | Brazil<br>(General population) | 104 | Mean = 73.6 | Clinical exam | 89.0 | 42.0 | [23] |
| PAR-Q (revised) | Brazil<br>(Elderly) | 89 | 61 – 89 | Clinical exam (absolute contraindications) | 75.0 | 19.8 | [38] |
|  |  |  |  | Clinical exam (relative contraindications) | 77.8 | 19.7 |  |
| Cornell Medical Index | Canada<br>(University staff) | 74 | 45-75 | Clinical exam | 35.0 | 92.6 | [50] |
| GAQ | Canada | 112 | Mean = 75.0 | Clinical exam, stress ECG | 6.0 | 95.0 | [4] |
| GAQ | Canada | 207 | 3 – 14 | Clinical exam / clinical notes | 71.0 | 59.0 | [33] |
| AAPQ | Brazil | 69 | 40 - 81 | Clinical exam, stress ECG | 88.2 | 13.5 | [34] |

*PAR-Q* – Physical Activity Readiness Questionnaire; *GAQ* – Get Active Questionnaire; *AAPQ* – American Heart Association/American College of Sports Medicine Pre-participation Questionnaire; *BP* – blood pressure; *ECG* – electrocardiogram.

The PAR-Q was revised in 1991 [54, 68] to improve clarity and address the high proportion of false positives from the question on blood pressure [49]. The revised PAR-Q [54] demonstrated reduced overall positivity versus the original [19, 21, 30] yet concerns remained regarding exclusions due to the question on blood pressure, particularly in older adults [18, 20, 48]. Until release of the PAR-Q+ and ePARmed-X in 2011, the PAR-Q was used by an estimated 50 million people [6]. In contrast, the PARmed-X was poorly adopted due to its length and the fact that some aspects of the form are based on opinion rather than being evidence-based [71]. By design, the PAR-Q and PARmed-X are purposely conservative, and concerns over the number of false positives existed from inception [3, 18, 49, 54]. A 2016 review estimated 10-30% of the general population were likely to be excluded by the PAR-Q, with a higher proportion in older adults [2]. The literature identified by our rapid review confirm these estimates (section 3.5, Table 2).

**Table 2.** – Proportion of pre-participation questionnaire respondents advised/referred to seek medical clearance.

| Screening Tool | Country (Setting) | Sample | Age | Positives (%) | Reference |
| --- | --- | --- | --- | --- | --- |
| PAR-Q<br>(19-item version) | Canada<br>(Agricultural exhibition) | 1,253 | Adults<br>(unspecified) | 52.4 | [70] |
| PAR-Q | Canada<br>(Insurance employees) | 1,130 | 18 - 70 | 20.0 | [49] |
| PAR-Q<br>PAR-Q (revised) | Canada | 399 | Adults<br>(unspecified) | 17.0<br>12.0 | [54] |
| PAR-Q<br>PAR-Q (revised) | U.S.A<br>(Community Nutrition Centres) | 169 | >70 | 82.3<br>62.1 | [19] |
| PAR-Q | U.S.A<br>(Masters athletes, general population) | 135 | 50 - 86 | 42.0 | [48] |
| PAR-Q<br>PAR-Q (revised) | U.S.A<br>(Community Nutrition Centres) | 193 | 60 - 69 | 75.7<br>66.3 | [21] |
| PAR-Q (revised) | U.S.A<br>(Community Nutrition Centres) | 181 | 60 – 89 | 54.7 | [18] |
| PAR-Q (revised) | U.S.A<br>(Community Nutrition Centres) | 200 | 60 – 79 | 77.5<br>64.0 | [20] |
| PAR-Q (revised) | USA<br>(Primary care) | 882 | Mean = 49.0 | 70.0 | [17] |
| PAR-Q<br>PAR-Q (revised) | U.K<br>(Leisure settings) | 50 | 16 – 69 | 36.0<br>32.0 | [30] |
| PAR-Q (revised) | Australia<br>(Primary care) | 330 | ≥ 65 | 33.3 | [51] |
| PAR-Q | Colombia<br>(Runners) | 445 | Mean = 37.7 | 16.2 | [24] |
| PAR-Q (revised) | Brazil<br>(Masters athletes) | 12 | Mean = 48.0 | 83.3 | [37] |
| PAR-Q (revised) | U.S.A<br>(NHANES 2001 – 2004) | 6,785 | $\geq 40$ | 68.4 | [55] |
| PAR-Q* | Brazil<br>(Leisure settings) | 50 | 18 – 35 | 20.0 | [12] |
| PAR-Q* | Colombia<br>(Runners) | 510 | 18 – 64 | 19.8 | [45] |
| PAR-Q* | Brazil<br>(Leisure settings) | 468 | 40 – 59 | 48.7 | [27] |
| PAR-Q* | Colombia<br>(General population) | 500 | $\geq 40$ | 64.0 | [42] |
| PAR-Q+ | Turkey<br>(Primary care) | 304 | Mean = 48.0 | 12.9 - 69.4<br>(Question specific) | [26] |
| PAR-Q+ & ePARmed-X | Canada | 1,670 | 18 – 66 | 0.5 – 14.7<br>(BP cut-offs) | [57] |
| PAR-Q+ (2015) | Japan<br>(Leisure settings) | 1,235 | 18 – 80 | 11.3 | [28] |
| PAR-Q* | Brazil<br>(Students) | 87 | Mean = 21.0 | 44.8 | [25] |
| PAR-Q (revised) | Spain<br>(Heart Failure patients) | 163 | Mean = 66.2 | 64.0 | [31] |
| PAR-Q+ | South Africa<br>(Runners, marathon) | 76,654 | Adults<br>(unspecified) | 23.2 | [52] |
| SMA-AAESS | Australia | 2,298 | 18 – 79 | 43.0 – 73.0 (M)<br>44.0 – 66.1 (F) | [39] |
| SMA-PESS | Australia<br>(Leisure centres) | 46 | Mean 40.0 | 54.0 | [16] |
| AHA | South Africa<br>(Runners, marathon) | 76,654 | Adults | 10.0 | [52] |
| AAPQ | U.S.A<br>(NHANES 2001 – 2004) | 6,785 | $\geq 40$ | 94.5 | [55] |
| ACSM 10<sup>th</sup> Edition | U.S.A<br>(NHANES 2001 – 2004) | 6,785 | ≥ 40 | 54.2 | [56] |
| ACSM 9<sup>th</sup> Edition<br>ACSM 10<sup>th</sup> Edition | South Africa<br>(Runners, marathon) | 76,654 | Adults | 33.9<br>6.7 | [52] |
| ACSM 9<sup>th</sup> Edition<br>ACSM 10<sup>th</sup> Edition | U.K<br>(Students) | 553 | Mean = 19.0 | 14.8<br>4.9 | [43] |
| EACPR | South Africa<br>(Runners, marathon) | 29,585 | Mean = 42.1 | 13.7 | [47] |
| EACPR | South Africa<br>(Runners, marathon) | 76,654 | Adults<br>(unspecified) | 33.9 | [52] |
| ESC/EACPR | South Africa<br>(Runners, marathon) | 15,778 | Adults<br>(unspecified) | 31.3 | [46] |

*\*Denotes unclear whether original or revised PAR-Q was used. PAR-Q – Physical Activity Readiness Questionnaire; SMA-AAESS – Sports Medicine Australia- Australian Association for Exercise and Sport Science; SMA-PESS – Sports Medicine Australia Pre-Exercise Screening System; AHA – American Heart Association; AAPQ – American Heart Association/American College of Sports Medicine Pre-participation Questionnaire; ACSM – American College of Sports Medicine; EACPR – European Association of Cardiovascular Prevention and Rehabilitation; ECR – European Society of Cardiology; NHANES – National Health And Nutrition Examination Survey.*

Barriers to exercise generated by the PAR-Q were addressed during design of the PAR-Q+ and ePARmed-X [6]. Collaboration between international authorities and fitness organisations began in 2007 to simplify pre-participation clearance using the Appraisal of Guidelines for Research and Evaluation (AGREE)[72] framework. After a series of systematic reviews, over 60 evidence-based recommendations of an expert consensus panel were incorporated into the 2011 PAR-Q+ and ePARmed-X [6]. During PAR-Q+ development, Canada gave university-educated fitness professionals greater responsibility to probe positive PAR-Q responses and provide clearance to exercise [61, 73] and suggested this model be adopted elsewhere.

The PAR-Q+ is a 4-page form designed to stratify an individual using an evidence-based risk continuum. Like the original PAR-Q, answering “No” to all Page 1 questions clears the individual for exercise, and provides generalised activity recommendations. One or more positive responses on Page 1 directs the individual to Pages 2 and 3 containing follow-up questions on various chronic conditions. Answering “No” to all follow-up questions on Pages 2 and 3 permits the individual to increase their activity. Answering “Yes” refers the individual to an exercise professional and/or the online ePARmed-X for additional probing questions [59], which guide the participant toward an appropriate intensity of exercise based on the risk continuum. Whilst the PAR-Q+ collaboration estimated physician referrals from the PAR-Q+/ePARmed-X to be 1% [74], there are a limited number of studies by independent researchers that replicate these estimates, and most suggest the proportion of referrals is higher (Table 2). The PAR-Q+ Collaboration regularly evaluate and update each document based on user feedback and research trials (www.eparmedx.com). However, the Get Active Questionnaire is now the CSEP-endorsed pre-screening tool [75], the ACSM have created their own screening guidelines based on a risk-stratification approach that considers exercise intensity to increase accessibility to low-to-moderate intensity exercise [76], and many UK organisations (UK Active, Everyone Active) have adopted a ‘Health Commitment Statement’ over the PAR-Q. According to UK Active [77], the Health Commitment Statement forms part of the contract between individuals and their fitness operator to deliver an appropriate balance of responsibility between parties. This approach is used in more than 1,000 facilities across the UK, and UK Active state that this “eliminates many of the flaws found in the PAR-Q and delivers a better outcome for everyone”. Whilst designed by industry experts and reviewed regularly for medicolegal integrity, it is appropriate to highlight that there is little peer-reviewed research on the Health Commitment Statement.

### 3.4 - What is the diagnostic accuracy of the PAR-Q? (Aim 1c)

**The PAR-Q and other pre-participation questionnaires lack the ability to accurately distinguish individuals with exercise contraindications from those at low risk**

*Table 1 contains sensitivity and specificity data for pre-participation questionnaires*

Validation of the PAR-Q was originally conducted in North America in the 1980s. The 19-item questionnaire was compared against (a) clinical examination plus blood pressure, (b) clinical examination plus blood pressure and resting ECG, (c) clinical examination plus blood pressure, resting ECG and stress ECG. Values of 78.5%/92.5% for sensitivity/specificity, respectively, were highest for clinical examination plus blood pressure but reduced to 41.8%/41.8% when both ECGs were included. The authors estimated the 7-item questionnaire would achieve 60-70% sensitivity and >80% specificity [70] but a subsequent validation report suggested lower sensitivity and higher specificity (36.7% and 95.5%, respectively) [22]. Another study of 1,130 sedentary office workers reported perfect sensitivity (100.0%) and high specificity (81.5%) against clinical examination, but the PAR-Q poorly identified those with stress ECG abnormalities (sensitivity = 33.3%, specificity = 82.1%) [49]. Specificity against clinical examination was similar in a sample of university staff (83.3%) with sensitivity lower than originally proposed estimates (55.0%)[50]. These studies appear to be the underpinning validation of the PAR-Q.

Validity of the revised PAR-Q [54] was later studied in two Brazilian samples of older adults. The first compared the tool against clinical examination with sensitivity of 89.0% and specificity of 42.0%, advocating poor accuracy in older adults [23]. Another used clinical examination to identify the ACSM’s [78] absolute and relative contraindications. Reasonable sensitivity was observed for absolute and relative contraindications (77.0% and 77.8%, respectively) with poor specificity (19.8% and 19.7%). Area under the curve (AUC) values of 0.47 and 0.49 for absolute and relative contraindications, respectively, indicated poor diagnostic power [38]. Question-specific analysis also reflected poor diagnostic power with AUC values of 0.40 to 0.53. The authors highlighted that “clinically very useful” tools should achieve 100% sensitivity and 80% specificity with AUC values ∼0.90, raising similar concerns over the PAR-Q in older adults [79]. Our rapid review did not identify any studies on the validity of the widely endorsed PAR-Q+/ePARmed-X.

Other pre-participation questionnaires do not appear to outperform the PAR-Q. High sensitivity (88.2%) and low specificity (13.5%) of the American Heart Association/ACSM’s AAPQ versus cardiology screening had low diagnostic capacity (AUC = 0.51) in another Brazilian sample [34]. In contrast, the CSEP’s Get Active Questionnaire (GAQ) had poor sensitivity (6.0%) but high specificity (95.0%) versus an exercise stress test in older adults, demonstrating good ability to clear those safe to exercise but poor ability to identify those at risk [4]. The GAQ performed slightly better in a sample of children where parents’ questionnaire responses were compared to medical records, with sensitivity of 71.0% and specificity of 59.0% [33]. The prohibitive nature of clinical examination and stress ECG must be judged against the limited accuracy of pre-participation questionnaires.

### 3.5 - What proportion of individuals are referred to seek medical clearance by pre-participation screening questionnaires? (Aim 2a)

**Original estimates suggested 1 in 5 PAR-Q respondents would be referred for medical clearance before increasing their physical activity. Subsequent studies suggest this is an underestimation, especially for older individuals**

*Table 2 contains the percentage of positive respondents for pre-participation questionnaires*

Considerably more publications report the percentage of individuals responding positively (i.e indicating they should be referred to seek medical clearance) than sensitivity and specificity. This is likely because it is easier to record positive responses than to compare questionnaire responses against clinical examination. Data exist from several populations and settings, with a broad range of sample sizes (*n* = 12 to *n* = 76,654, Table 2).

The original 19-item PAR-Q led to positive responses in 52.4% of participants [70]. The authors suggested the reduced 7-item version would lead to 15-25% positivity [22]. Whilst this reduced positivity was confirmed (20%) in a subsequent study, the authors questioned if such an exclusion rate was acceptable [49]. Each of these studies included a range of adult age groups. Notably higher positive responses are evident in studies of older adults, ranging from 42.0% to 82.3% [19, 21, 48]. High positive responses to the question on blood pressure [48, 49] prompted work to revise the PAR-Q. Reduced positivity in the revised version from 17.0% to 12.0% included fewer positive responses to the blood pressure question [54], and studies directly comparing the original and revised PAR-Q verified a reduction in overall positivity [19–21, 30]. However, positivity of the revised PAR-Q in the available literature ranges considerably from 12.0% to 83.3%, with higher values in older cohorts persisting [17–21, 31, 37, 51, 55]. Despite revisions, the blood pressure question still accounted for a higher percentage of positive responses than other items [18, 20, 48]. With some exceptions, most published data on positive response rates of the original and revised PAR-Q are higher than the originally estimated 15-25%, and considerably higher in participants over 40. This remains in cohorts of active older adults [27, 37, 48], suggesting age drives positivity more than activity level.

Retrospective application of the revised PAR-Q to the National Health and Nutrition Examination Survey (NHANES) suggested 68.4% of Americans aged over 40 would be referred for medical clearance [55]. By comparison, the AAPQ identified more (94.5%) and the ACSM fewer (54.2%) when applied to the same sample [56]. A recent investigation concluded that the revised PAR-Q and PAR-Q+ would fail to detect 52.5% and 45.5% of medical abnormalities, respectively, with both performing poorly compared to versions of the AAPQ (9.7% - 16.6%) and EACPR (4.4% - 17.3%) [41]. A retrospective study of 76,654 endurance race entrants suggested between 6.9% and 33.9% would require medical clearance based on their responses to a pre-race questionnaire [52]. The PAR-Q+ would have excluded 23.2%, which was more conservative than the most recent ACSM guidelines (6.7%) and the AHA questionnaire (10.0%), but less than earlier ACSM guidelines and the EACPR questionnaire (both 33.9%). Another study combined the PAR-Q+ with blood pressure readings to investigate how different blood pressure cut-offs impacted exclusion [57]. The most conservative cut-off of <130/80 mmHg led to 14.7% exclusions, reducing incrementally until 0.5% at the <160/100 mmHg cut-off. Those with blood pressure >160/90 mmHg were further screened via ePARmed-X and all were cleared for maximal exercise. Finally, a 2016 conference abstract describes PAR-Q+ as a follow-up to risk stratification based on ACSM guidelines. All “high-risk” college students requiring medical clearance (ACSM 9^th^ edition = 16.9%, ACSM 10^th^ edition = 9.5%) were cleared for sub-maximal activity via PAR-Q+/ePARmed-X and completed exercise testing [15].

Beyond those referenced above, a small number of studies describe other questionnaires. Applied retrospectively, the EACPR would have excluded 13.7% of South Africans from competing in an endurance event [47], whereas combined guidelines from the EAPCR and ECR would have excluded 31.3% [46]. A UK-based study in students demonstrated that changes in the ACSM guidelines between the 9^th^ and 10^th^ editions reduced positivity from 14.8% to 4.9% [43]. Versions of the Sports Medicine Australia guidelines would require 43-73% of male and 44-61% of female survey respondents (SMA-AAESS) [39] or 54.0% of fitness centre members (SMA-PESS) [16] to seek clearance.

### 3.6 - Is there any associated impact on mortality and/or adverse events? (Aim 2b)

**Evidence is very limited in both quantity and quality regarding an association of pre-participation questionnaires with subsequent mortality and exercise-related adverse events**

Limited evidence exists linking PAR-Q response with mortality or exercise-related adverse events. A 7-year follow-up study of 31,668 Canadian citizens aged 15-64 found higher morbidity and mortality in persons responding positively to the original PAR-Q. Specifically, they observed 2.2 higher odds for all-cause mortality and 9.1 higher odds for death from cardiovascular disease versus those with no positive answers [13]. Two earlier mentioned studies observed no adverse events during or post-exercise using the PAR-Q+/ePARmed-X: the first cleared 1,670 participants screened using PAR-Q+/ePARmed-X and blood pressure, despite a sufficient number with pre-hypertensive or hypertensive readings [57]; the second cleared “high-risk” students for exercise testing and observed no adverse events [15]. A recent pre-print reported no adverse events or near-misses in 1,235 Japanese gym users during a 14-month study period, though it is not clear if the 11.3% who responded positively to the PAR-Q+ were cleared to exercise [28]. In another study of 29,585 ultra-marathoners, exercise-related adverse events were significantly higher in those deemed “very high risk” using a pre-participation questionnaire based upon EACPR guidelines [47]. Logistical challenges of longitudinal investigations, either prospective or retrospective, offer plausible explanation for limited evidence in this area.

### 3.7 - What are the limitations of pre-participation questionnaires? (Aim 1d)

**Key limitations of pre-participation questionnaires include not being evidence-based, the binary nature of questions, and a lack of agreement with physician-led medical screening**

Several limitations of pre-participation questionnaires exist. The opinion-based nature of the original and revised PAR-Q is a key limitation [2]. Despite adopting the AGREE process and accepting recommendations based on systematic reviews, the newer PAR-Q+ and ePARmed-X are also consensus-based. This is a similar limitation of the AHA questionnaire and ACSM guidelines [2], and is underpinned by a lack of up-to-date validation studies. Importantly, PAR-Q+ and ePARmed-X clearances are applicable for 12 and 6 months, respectively [59], suggesting the tool is only valid if regular screening is employed to detect changes in health status.

The limitations of any tool include variability in adherence, application and/or interpretation. Indeed, reports suggest a significant number of facilities do not perform pre-participation screening [36, 80]. One report demonstrated higher positivity of both the original and revised PAR-Q in the UK versus the USA and Canada [30], suggesting geographical inconsistencies in how the same questionnaire is administered to similar populations. Prior to the PAR-Q+, final clearance relied on the decision of a physician. A study found that Israeli physicians who exercised themselves were more likely to grant clearance to an individual responding positively on the PAR-Q than inactive colleagues [29], which could question the use of clinician sign-off as a gold standard for validation studies.

The dichotomous “Yes/No” nature of many screening tools, particularly the PAR-Q, lacks context and likely contributes to the high number of positive responses. Whilst the PARmed-X was designed to help physicians apply further context to this scenario, it was poorly adopted. The PAR-Q+ and ePARmed-X aim to involve qualified exercise professionals in screening and risk stratification to reduce unnecessary barriers to acivity [6]. Consequently, stratification-based tools like the PAR-Q+/ePARmed-X and the most recent ACSM guidelines [76] are likely to reduce referral rates through a more informed process. This assumes that the PAR-Q+/ePARmed-X are widely adopted, which is hard to establish from published literature. Nevertheless, some evidence suggests that the PAR-Q+/ePARmed-X might remove barriers to exercise in higher risk individuals if a qualified exercise professional is involved in screening [15, 57]. However, not all countries have a recognised exercise professional qualification, which may limit the ability of fitness staff to apply the PAR-Q+ properly.

Data on the number of positive responses (Table 2) suggests pre-participation questionnaires will refer 1 in 5 participants for medical screening, increasing with age regardless of activity level. Whilst replicated most in studies of the original/revised PAR-Q, similar patterns exist for the ACSM guidelines (9^th^ and 10^th^ edition), AAPQ, EACPR, and questionnaires by Sports Medicine Australia. This is particularly problematic considering increases in life expectancy and the number of older adults seeking the benefits of physical activity. Such high rates of referral/exclusion undoubtedly arise from a high number of false positives, as suggested by specificity data (Table 1). Indeed, the original PAR-Q validation studies reported 41.2% false positives for the 19-item version [70] and 19.0% for the 7-item version [49]. High false positive rates were also a concern for the revised PAR-Q [38], AAPQ [34], and GAQ [33]. False positives in those who stand to gain the most from physical activity, such as individuals who are inactive and those with multimorbidity [6], present an unacceptable barrier to exercise. Comparatively, low sensitivity of a pre-participation questionnaire can lead to “at-risk” participants being cleared to exercise, as demonstrated when screening tools fail to predict ECG abnormalities observed under exercise conditions [22, 49, 65]. False negatives due to low sensitivity also raise concern because missed contraindications place participants at undue risk.

### 3.8 – Limitations of this rapid review

A rapid review was considered appropriate to expedite information synthesis with the time and resources available. Nonetheless, the use of restricted methods incurs limitations that must be acknowledged. We were unable to involve an information specialist when developing the search strategy. Consequently, some sources of information may have been overlooked. Peer review of the search strategy using the PRESS checklist was not conducted. However, the search strategy was double checked independently by two co-authors for typographical errors, missed key words, and correct use of Boolean operators, thus improving the rigour of the search strategy. Due to time and resource constraints, we were unable to pilot and refine processes for article screening and data extraction, nor was it possible to complete dual and independent screening, asses risk of bias, or grade certainty of the evidence extracted. It is acknowledged that adhering to these recommendations increases objectivity and reduces the risk of bias, which should be considered when interpreting the conclusions of this review.

## 4. Conclusions

The evidence identified in this rapid review suggests there are several limitations to pre-participation questionnaires. A high number of false positives creates unnecessary barriers to exercise, particularly in older adults. In contrast, sub-optimal test sensitivity increases the risk of “at-risk” individuals being placed at undue risk.

Key points from this rapid review are:

- The PAR-Q was the original international standard pre-participation questionnaire. Versions of the PAR-Q are most common in scientific studies.
- The PAR-Q was developed in Canada in the 1980s during design of the Canadian Home Fitness Test and was originally validated on conference attendees and sedentary office workers.
- The key limitations of pre-participation questionnaires are poor agreement with medical examination and a high number of people referred to seek medical clearance.
- Pre-participation questionnaires refer approximately 1 in 5 people to seek medical clearance before engaging in exercise. This is considerably higher in older individuals.
- Evidence linking pre-participation questionnaires with subsequent mortality, and the ability to predict/reduce exercise-related adverse events, is extremely limited.

## 5. Summary

Concerns exist around inactive people starting physical activity, particularly if they have medical conditions. The main worry is that being active could cause a new problem, for instance with their heart. However, increasing their activity levels is very important for improving their health in the long term. A range of screening tests and questionnaires have been developed to try and help identify who is safe to exercise and who needs a medical review first. Despite a great deal of work going in to developing and improving the questionnaires, they are not reliable at both identifying who is at high risk of a medical problem and needs medical attention nor do they accurately state who is at low risk and can carry on without medical review. Being able to answer both questions accurately is important for physical activity organisations such as gyms and leisure centres as well as members of the public. Further work is needed to improve this process.

## Data Availability

All data produced in the present work are contained in the manuscript

## Notes

### Competing Interest Statement

The authors have declared no competing interest.

### Funding Statement

This study did not receive any funding

